# Results of the inoperable and operable with aortic valve endocarditis

**DOI:** 10.1101/2023.09.06.23295165

**Authors:** Jing-bin Huang, Zhen-zong Du, Chang-chao Lu, Jian-rong Yang, Jun-jun Li

## Abstract

**Objectives:** To evaluate the results of the inoperable and operable with aortic valve endocarditis.

**Methods:** This was a retrospective study of patients with aortic valve endocarditis undergoing cardiac surgery between January 2006 and November 2022 at our hospital.

**Results:** 512 patients were divided into group with destruction of the aortic annulus (n=80) and without destruction of the aortic annulus (n=432). There were 32 operative deaths (6.3%, 32/512). Univariate and multivariate analysis showed that destruction of the aortic annulus is statistically significantly associated with in-hospital mortality (P<0.001), prolonged mechanical ventilation time (mechanical ventilation time> 96h, P=0.018), early aortic paravalvular leak (P<0.001), and 1-year mortality following cardiac surgery (P<0.001), respectively.

**Conclusions:** In our study, destruction of the aortic annulus increases mortality and health care costs. Optimization of pre-, peri-, and postoperative factors can reduce mortality and morbidity in aortic valve endocarditis. Aortic root replacement could be recommended as the best practice choice for aortic valve endocarditis with periannular abscess and destruction of the aortic annulus.

## Introduction

Infective endocarditis (IE) is defined by a focus of infection within the heart and is a feared disease across the field of cardiology. Even at experienced centers, operations for IE remain associated with the highest mortality of any valve disease. Approximately half of all IE patients are identified as high-risk and undergo operative treatment. Destruction of the aortic annulus, which is caused by aortic periannular abscess or aortic root abscess, is a severe complication of aortic valve endocarditis. Surgical treatment of aortic valve endocarditis with destruction of the aortic annulus is a challenging issue with high mortality and morbidity rate in the current era. [1–3]

In our cohort, the main reasons for perioperative death were paravalvular leaks and septic shock with consecutive multiorgan failure and cerebral hemorrhage. One of the most frequent determinants of paravalvular leaks is destruction of the aortic annulus in aortic valve endocarditis. However, only a few reports in the available literature evaluated the incidence and predictors for destruction of the aortic annulus in aortic valve endocarditis.[4] We aimed to investigate risk factors and significance of destruction of the aortic annulus in aortic valve endocarditis.

We hypothesized that optimization of pre-, peri-, and postoperative factors can reduce mortality and morbidity in aortic valve endocarditis.

## Patients and Methods

### 1.1 Design

This was a retrospective study of patients with aortic valve endocarditis undergoing cardiac surgery between January 2006 and November 2022 at our hospital. Medical records were reviewed.

### 1.2 Diagnosis

Patients were diagnosed according to the modified Duke criteria. [5] Surgical and pathological findings were reviewed to confirm the preoperative diagnosis.

### 1.3 Eligibility criteria

Inclusion criteria included patients with aortic valve endocarditis undergoing cardiac surgery between January 2006 and November 2022 at our hospital. Exclusion criteria included patients without aortic valve endocarditis. (Figure 1)

**Fig. 1.**
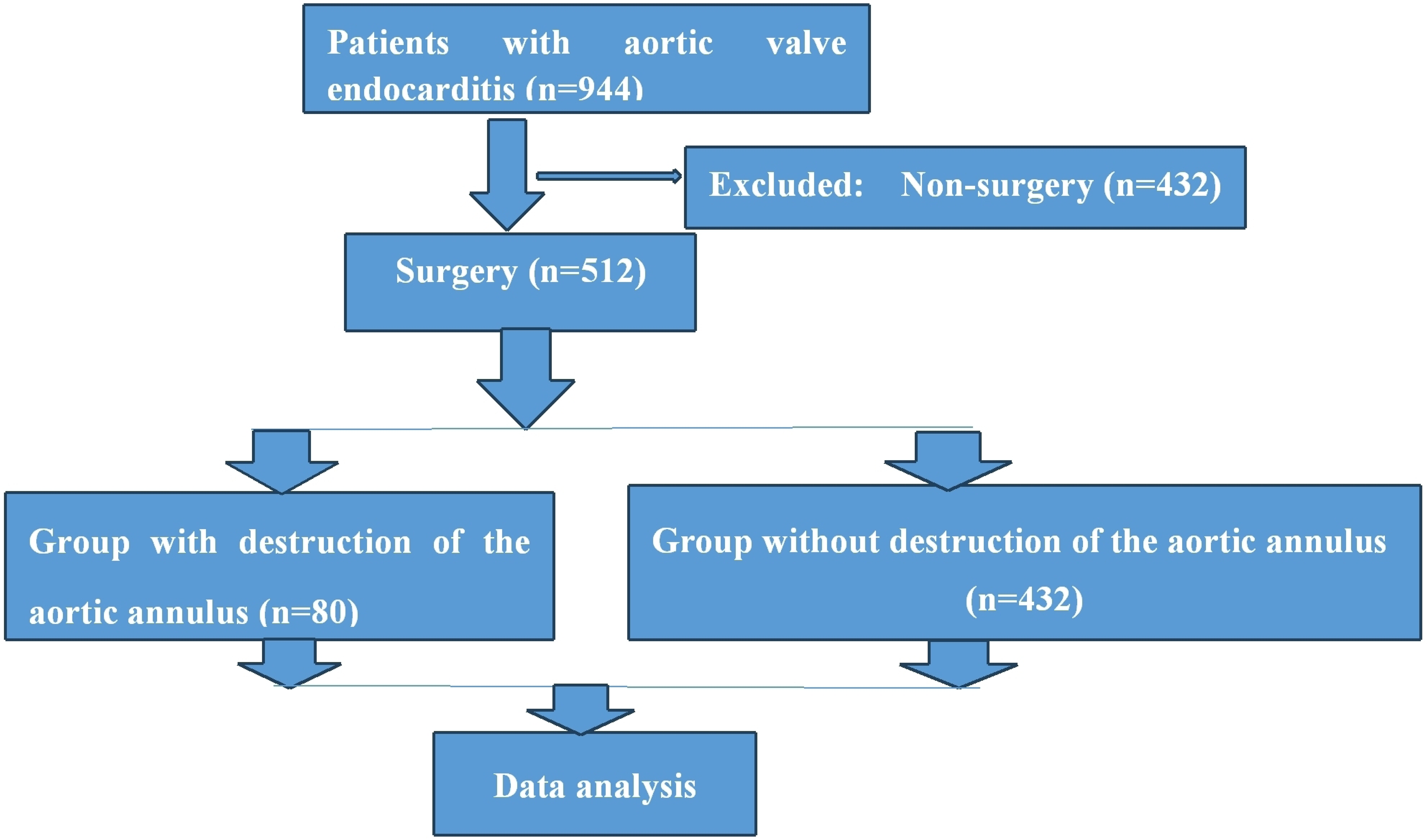
Flow chart of clinical trial

### 1.4 Surgical technique

Cardiac surgery for IE was indicated according to AATS guidelines for the management of IE. Radical debridement of all infected tissues and foreign material is followed by generous irrigation. Allografts are preferred for aortic root reconstruction in patients with annulus destruction.

### 1.5 Peri- and postoperative management

All patients were transferred to the intensive care unit after the operation. Anticoagulation management was conducted routinely. Transthoracic echocardiograph was performed postoperatively 1 to 7 days in intensive care unit.

### 1.6 Variables to be analyzed

Variables were evaluated. (Supplementary material)

Time between symptoms and admission was defined as length from onset of symptoms to the date of admission.

Time between symptoms and surgery was defined as length from onset of symptoms to the date of operation.

In-hospital mortality was defined as death within 30 days of the operation or during the same hospital admission.

In our study, serum creatinine was used as the diagnostic standard of acute renal injury. According to Kidney Disease Improving Global Outcomes (KDIGO) classification, if serum creatinine increases by ≥ 0.3 mg/dl (26.5 μmol/l) within 48 hours, serum creatinine is 50% higher than the baseline within first seven days, or urine output is below 0.5 ml/kg/hour for six hours, the patient is considered to have acute renal injury.[5]

Multiorgan failure (MOF) is regarded as a continuous process of varying levels of organ failure rather than an all-or-none event. To characterize MOF, six different organ systems are regarded as “key organs”: lungs, cardiovascular system, kidneys, liver, coagulation system, and central nervous system.[6]

Hepatic failure is defined as a severe liver injury, potentially reversible in nature and with onset of hepatic encephalopathy within 8 weeks of the first symptoms in the absence of pre-existing liver disease.[7]

Respiratory failure is a condition in which the respiratory system fails in one or both of its gas exchange functions, i.e., oxygenation of and/or elimination of carbon dioxide from mixed venous blood. It is defined by an arterial oxygen tension (Pa, O2) of ≤ 8.0 kPa (60 mmHg), an arterial carbon dioxide tension (Pa, CO2) of ≥6.0 kPa (45 mmHg) or both.[8]

### 1.7 Follow-up

All survivors discharged from hospital were monitored to the end date of the study. At the outpatient department, all patients were investigated with echocardiogram, electrocardiogram, and X-ray chest film, once every 3 to 12 months. At the last follow-up, the patients were contacted by telephone or micro-message or interviewed directly at the outpatient department.

### 1.8 Statistical analyses

Continuous variables are reported as means ± SE. Survival rates were estimated using the Kaplan-Meier method. The chi-square test, the Kruskal-Walls test or the Wilcoxon rank-sum test, as appropriate, to be used to evaluate relationships between the preoperative variables, and selected intraoperative and postoperative variables. The relationships with perioperative risk factors were assessed by means of contingency table methods and logistic regression analysis. P values less than 0.05 were statistically significant. All analyses were performed using IBM SPSS version 24.0 software (IBM SPSS Inc., USA).

## 2. Results

### 2.1 Multiorgan failure at admission in aortic valve endocarditis

During the study period, 944 patients were diagnosed as aortic valve endocarditis, with 272 (28.8%, 272/944) excluded from surgery because of multiorgan failure at admission, and 512 (54.2%, 512/944) undergoing surgery.

Body weight (58.97±0.28 versus 55.14±0.45 kg, P< 0.001), time between symptoms and admission (3.09±0.13 versus 2.32±0.09 months, P<0.001), vegetation length (14.42±0.29 versus 10.26±0.23 mm, P<0.001), aortic insufficiency (8.86±0.53 versus 5.68±0.24 cm^2^, P<0.001), and symptomatic neurological complications (47.1 versus 11.9 %, P < 0.001) in group of multiorgan failure at admission were significantly higher than those in group of non-multiorgan failure. (Table 1)

**Table 1.** Multiorgan failure at admission in aortic valve endocarditis.

| Variable | Group of multiorgan failure (n=272) | Group of non-multiorgan failure (n=672) | P value |
| --- | --- | --- | --- |
| Male | 208 (76.5%) | 512 (76.2%) | 0.927 |
| Age | 40.29±0.71 | 42.31±0.6 | 0.054 |
| Body weight | 58.97±0.28 | 55.14±0.45 | <0.001 |
| Time between symptoms and admission | 3.09±0.13 | 2.32±0.09 | <0.001 |
| Vegetation length | 14.42±0.29 | 10.26±0.23 | <0.001 |
| Aortic insufficiency | 8.86±0.53 | 5.68±0.24 | <0.001 |
| Symptomatic neurological complications | 128 (47.1%) | 80 (11.9%) | <0.001 |

### 2.2 Analysis of risk factors of multiorgan failure at admission in aortic valve endocarditis (n=432)

Univariate analysis showed that factors are associated with multiorgan failure at admission, including body weight (P<0.001), time between symptoms and admission (P=0.037), vegetation length (P < 0.001), aortic insufficiency (P < 0.001), and symptomatic neurological complications (P<0.001).

When they were included in multivariate analysis models, multivariate analyses also showed that factors are associated with multiorgan failure at admission, including body weight (P < 0.001), time between symptoms and admission (P < 0.001), vegetation length (P<0.001), aortic insufficiency (P<0.001), and symptomatic neurological complications (P<0.001). (Table 2)

**Table 2.** Analysis of risk factors for multiorgan failure at admission in aortic valve endocarditis (n=272)

| Model | OR | 95% CI | P value |
| --- | --- | --- | --- |
| Univariate analysis |  |  |  |
| Body weight | 0.960 | 0.945-0.975 | <0.001 |
| Time between symptoms and admission | 0.868 | 0.817-0.922 | <0.001 |
| Vegetation length | 0.891 | 0.869-0.914 | <0.001 |
| Aortic insufficiency | 1.059 | 1.033-1.084 | <0.001 |
| Symptomatic neurological complications | 6.578 | 4.713-9.181 | <0.001 |
| Multivariate analysis |  |  |  |
| Body weight | 0.955 | 0.937-0.974 | <0.001 |
| Time between symptoms and admission | 0.833 | 0.782-0.888 | <0.001 |
| Vegetation length | 0.890 | 0.867-0.914 | <0.001 |
| Aortic insufficiency | 1.080 | 1.052-1.109 | <0.001 |
| Symptomatic neurological complications | 7.610 | 5.145-11.256 | <0.001 |

### 2.3 Operative data

512 patients with aortic valve endocarditis undergoing cardiac surgery were divided into group with destruction of the aortic annulus (n=80) and without destruction of the aortic annulus (n=432). There were 32 operative deaths (6.3%, 32/512). (Table 3 and Table 4)

**Table 3.**
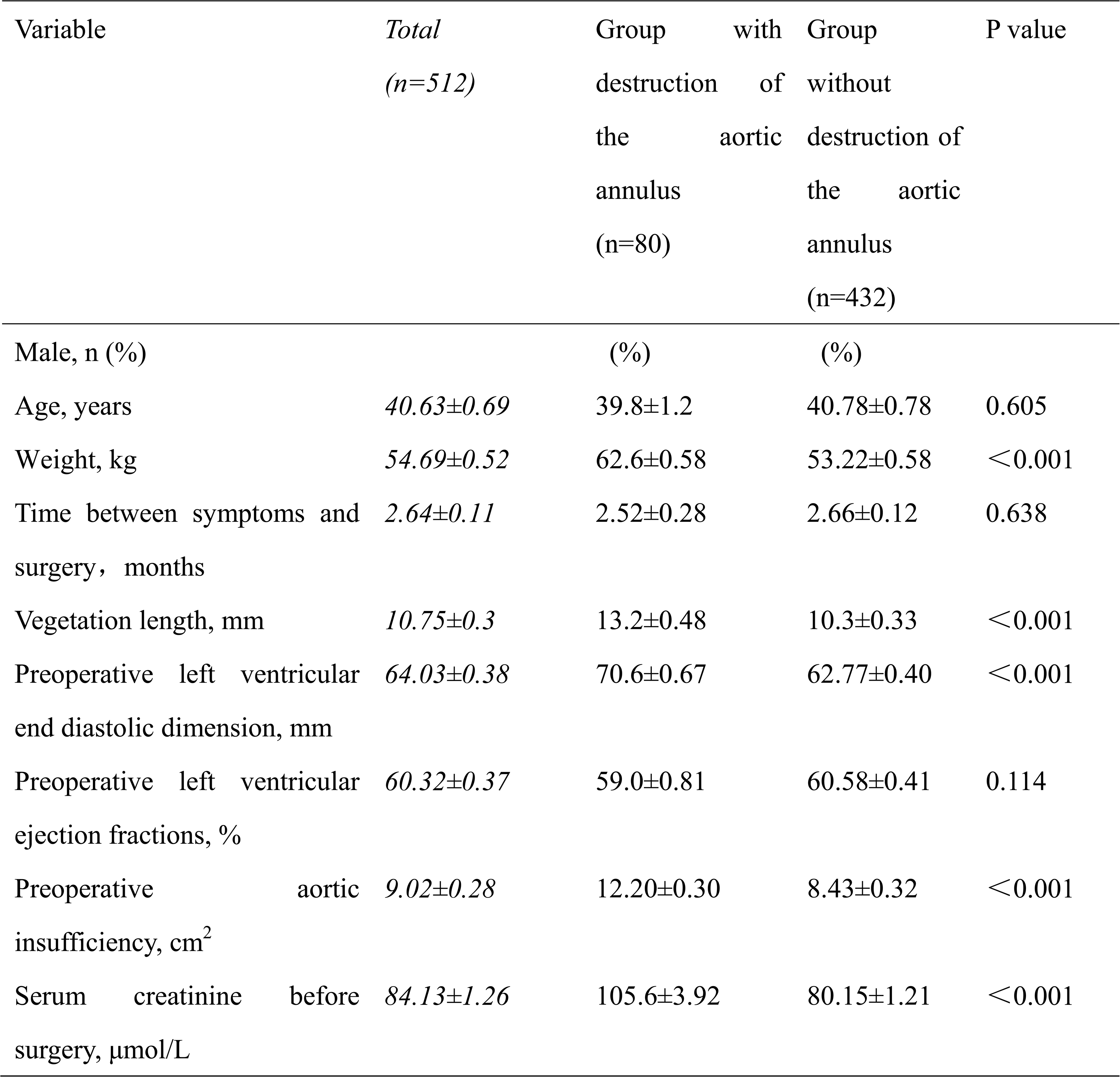
Preoperative data (n=512)

**Table 4.**
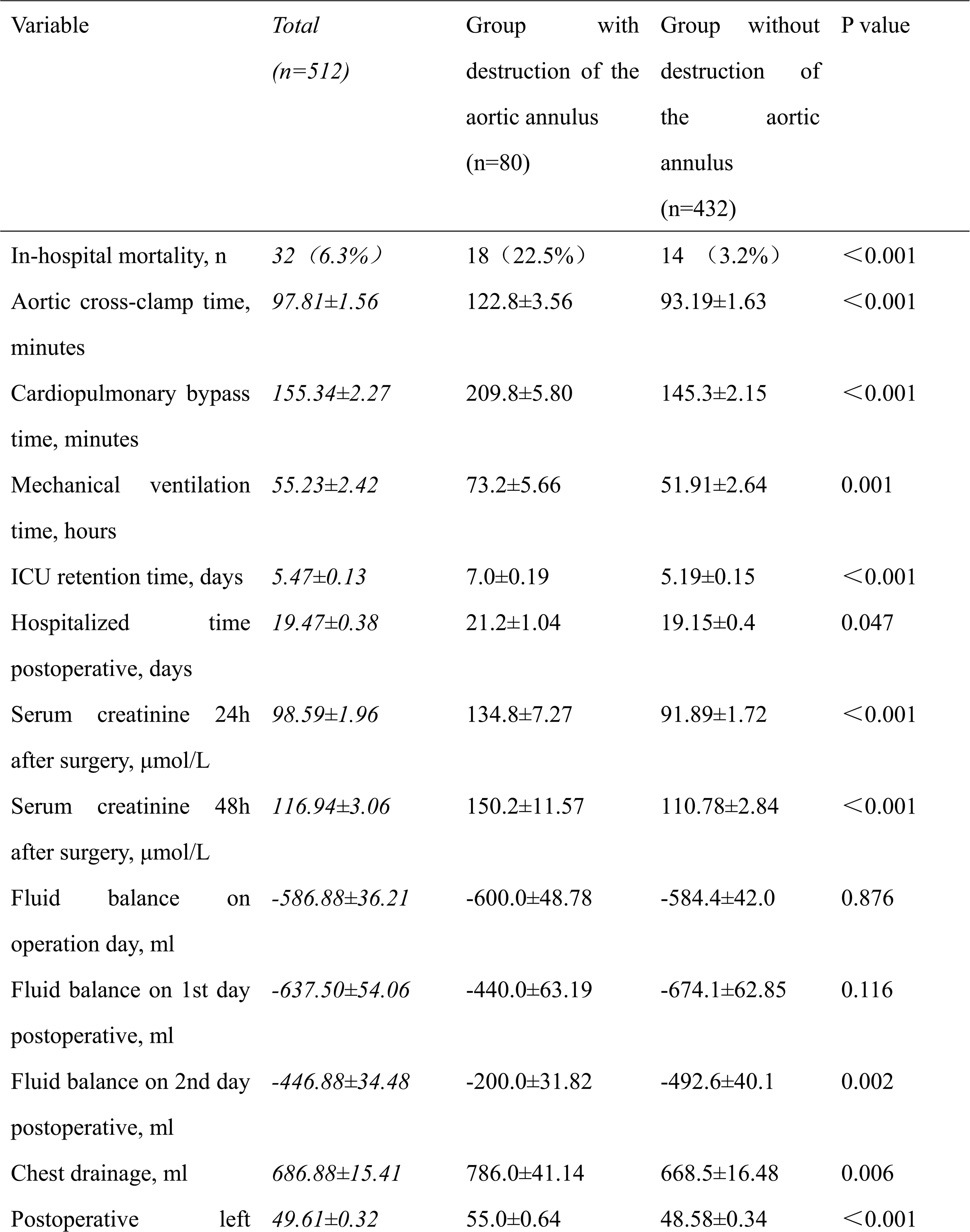
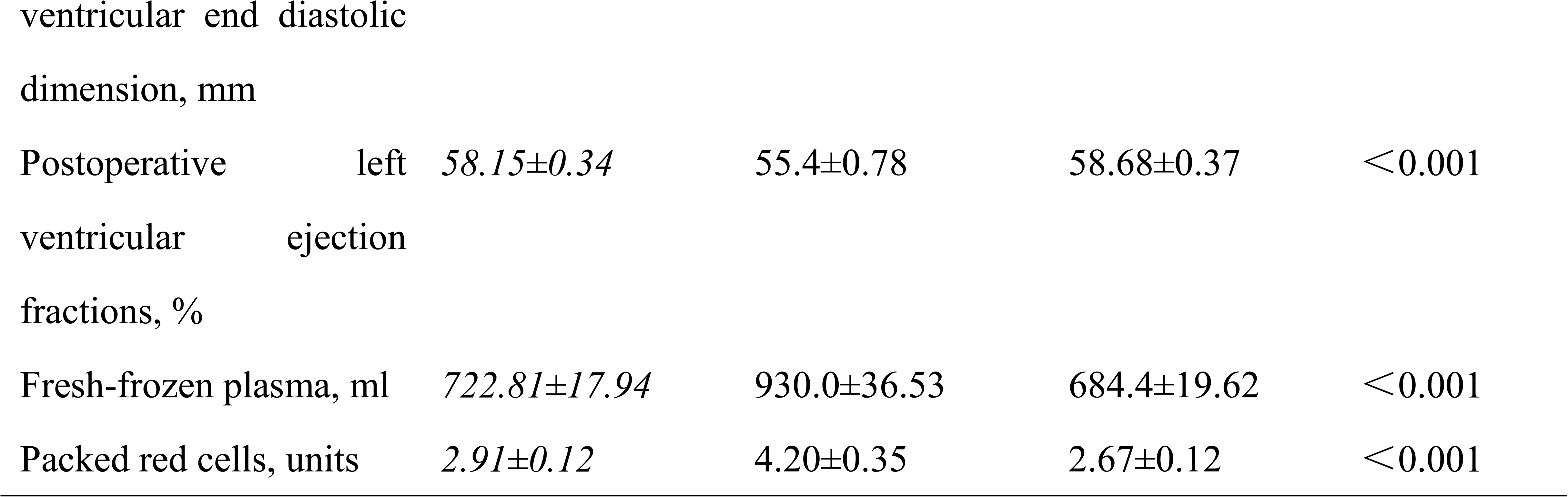
Operative data (n=512)

Weight (62.6±0.58 versus 53.22±0.58 kg, P < 0.001), vegetation length (13.2±0.48 versus 10.3±0.33 mm, P < 0.001), preoperative left ventricular end diastolic dimension (70.6±0.67 versus 62.77±0.40 mm, P<0.001), preoperative aortic insufficiency (12.2±0.30 versus 8.43±0.32 cm^2^, P<0.001), serum creatinine before surgery (105.6±3.92 versus 80.15±1.21 μmol/L, P<0.001) in group with destruction of the aortic annulus were significantly higher than those in group without destruction of the aortic annulus. (Table 3 and Table 4)

## Resource utilization

Aortic cross-clamp time (122.8±3.56 versus 93.19±1.63 minutes, P<0.001), cardiopulmonary bypass time (209.8±5.8 versus 145.3±2.15 minutes, P<0.001), mechanical ventilation time (73.2±5.66 versus 51.91±2.64 hours, P<0.001), ICU retention time (7.0±0.19 versus 5.19±0.15 days, P < 0.001), hospitalized time postoperative (21.2±1.04 versus 19.15±0.4 days, P=0.047), serum creatinine 24h after surgery (134.8±7.27 versus 91.89±1.72 μmol/L, P<0.001), serum creatinine 48h after surgery (150.2±11.6 versus 110.78±2.84 μmol/L, P<0.001), chest drainage (786.0±41.14 versus 668.5±16.48 ml, P=0.006), postoperative LVEDD (55.0±0.64 versus 48.58±0.34 mm, P < 0.001), fresh-frozen plasma (930.0±36.5 versus 684.4±19.62 ml, P<0.001), Fresh-frozen plasma (930.0±36.53 versus 684.4±19.62 ml, P<0.001), and packed red cells (4.2±0.35 versus 2.67±0.12 units, P<0.001) in group with destruction of the aortic annulus were significantly higher than those in group without destruction of the aortic annulus.(Table 4)

Fluid balance on 1st day postoperative (-440.0±63.19 versus -674.1±62.85 ml, P <0.001) and fluid balance on 2nd day (-200.0±31.82 versus -492.6±40.1 ml, P< 0.001) postoperative in group with destruction of the aortic annulus was significantly less negative than that in group without destruction of the aortic annulus.(Table 4)

The common early postoperative complications included acute renal injury (159/512, 31.1%), long-term intubation time>48h (230/512, 44.9%), and multiorgan failure (52/512, 10.2%).

### 2.4 Analysis of risk factors for destruction of the aortic annulus

Univariate analysis of potential risk factors for destruction of the aortic annulus in patients with aortic valve endocarditis showed that numerous factors are associated with destruction of the aortic annulus, including body weight (P<0.001), vegetation length (P<0.001), preoperative aortic insufficiency (P<0.001), preoperative left ventricular end diastolic dimension (P<0.001), and serum creatinine before surgery (P<0.001).

When they were included in multivariate analysis models, multivariate analyses also showed that numerous factors are associated with destruction of the aortic annulus, including body weight (P < 0.001), vegetation length (P < 0.001), preoperative aortic insufficiency (P < 0.001), preoperative left ventricular end diastolic dimension (P<0.001), and serum creatinine before surgery (P<0.001). (Table 5)

**Table 5.** Analysis of risk factors for destruction of the aortic annulus in aortic valve endocarditis.

| Model |  | OR | 95% CI | P value |
| --- | --- | --- | --- | --- |
| Univariate analysis |  |  |  |  |
| Body weight |  | 0.898 | 0.868-0.928 | <0.001 |
| Vegetation length |  | 0.942 | 0.911-0.974 | <0.001 |
| Preoperative aortic insufficiency |  | 0.919 | 0.888-0.952 | <0.001 |
| Preoperative left ventricular end diastolic dimension |  | 0.900 | 0.874-0.926 | <0.001 |
| Serum creatinine before surgery |  | 0.970 | 0.962-0.979 | <0.001 |
| Multivariate analysis |  |  |  |  |
| Body weight |  | 0.897 | 0.862-0.933 | <0.001 |
| Vegetation length |  | 0.926 | 0.887-0.967 | 0.001 |
| Preoperative aortic regurgitation |  | 0.934 | 0.897-0.972 | 0.001 |
| Preoperative left ventricular end diastolic dimension |  | 0.928 | 0.897-0.959 | <0.001 |
| Serum creatinine before surgery |  | 0.976 | 0.966-0.987 | <0.001 |

### 2.5 Analysis of significance of destruction of the aortic annulus in aortic valve endocarditis

Univariate and multivariate analysis of risk factors of in-hospital mortality, prolonged mechanical ventilation time (mechanical ventilation time> 96h), prolonged ICU retention time, early aortic paravalvular leak following cardiac surgery, and 1-year mortality following cardiac surgery in aortic valve endocarditis showed that destruction of the aortic annulus is statistically significantly associated with in-hospital mortality (P<0.001), prolonged mechanical ventilation time (mechanical ventilation time> 96h, P=0.018), early aortic paravalvular leak (P<0.001), and 1-year mortality following cardiac surgery (P<0.001), respectively. (Table 6)

**Table 6.**
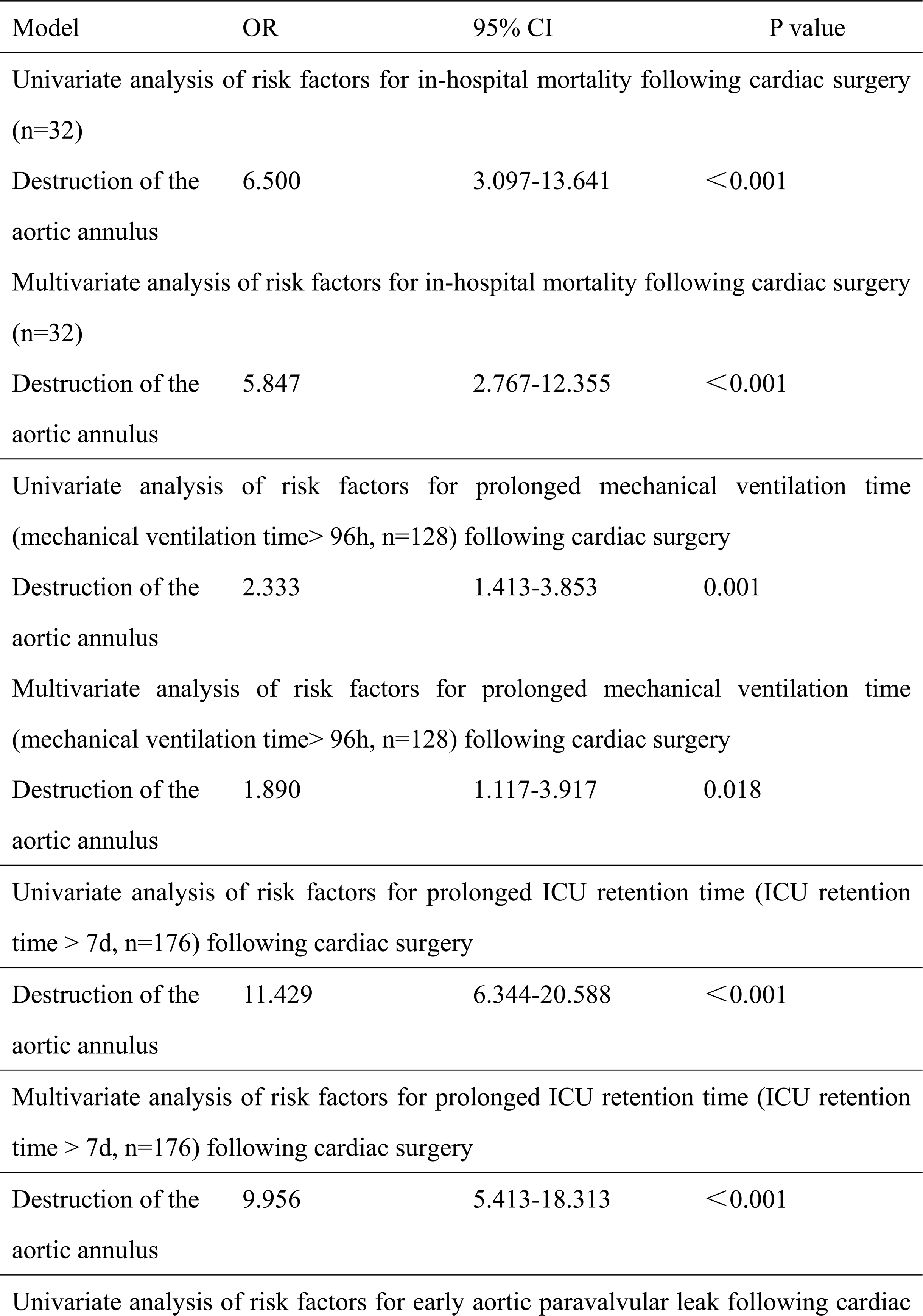
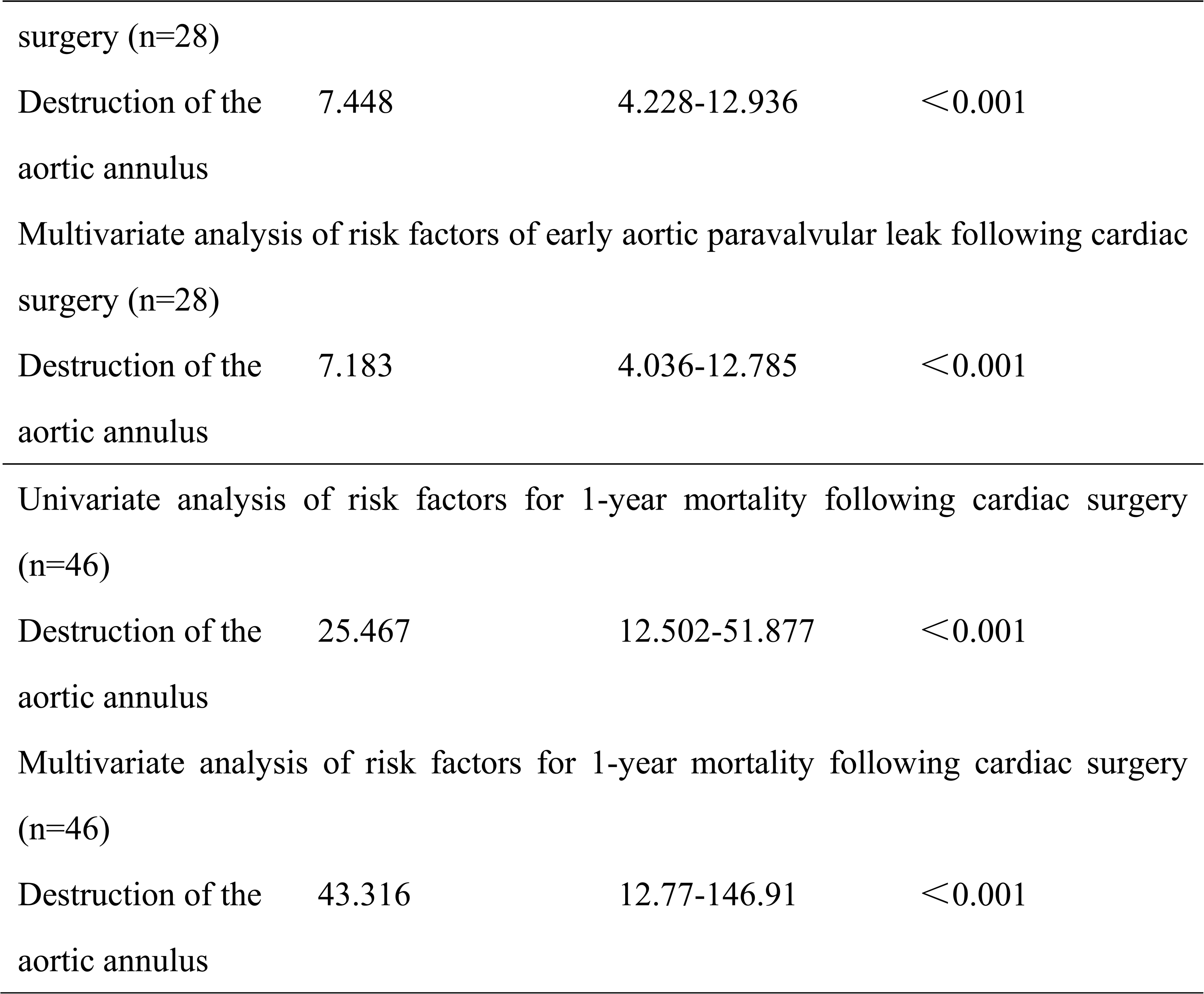
Analysis of the significance of destruction of the aortic annulus in aortic valve endocarditis (n=512)

### 2.6 Follow-up results

480 survivors were discharged and 460 patients were monitored to the end date of the study and the follow-up was 95.8% (460/480) completed. The mean duration of follow-up was 74.96 ± 2.44 months. 55 deaths (55/460, 12.0%) occurred within 12 months after being discharged because of recurrence of IE and cerebral hemorrhage. The latest data of follow-up showed that 373 survivors were in NYHA class I (373/405, 92.1%) and 32 in class II (32/405, 7.9%). (Figure 2)

**Figure 2.**
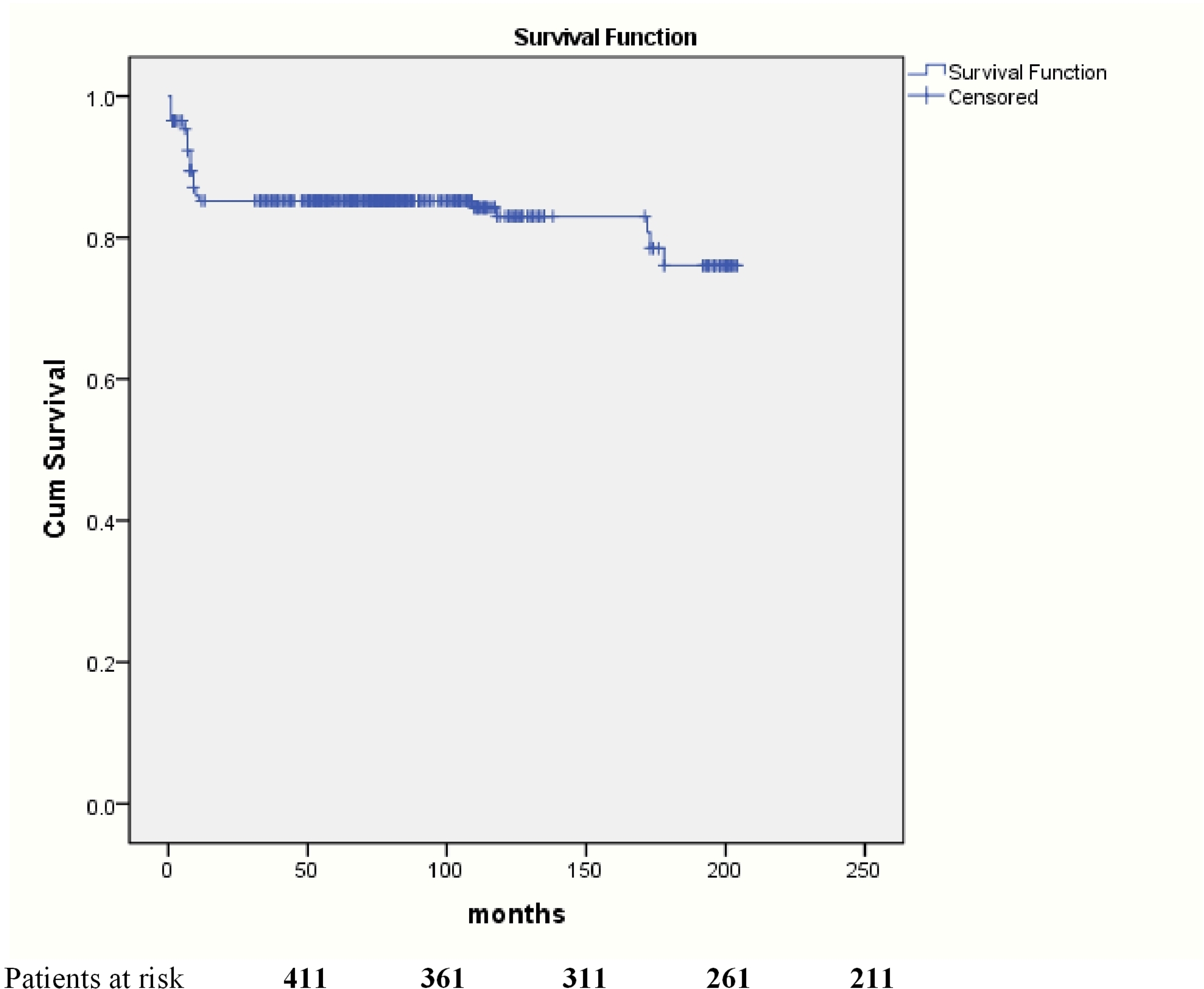
**Kaplan-Meier curve for survival.**

## 3. Discussion

### Destruction of the aortic annulus increases mortality and health care costs

The presence of a paravalvular abscess or destruction of aortic annulus is a crucial indication for surgical management due to its known association with high in-hospital mortality. Periannular abscess or destruction of aortic annulus occurs in more than 10% of patients with aortic valve endocarditis. The previous study indicated that about 22% of patients who suffered from aortic valve endocarditis had periannular abscess and destruction of aortic annulus. Another study reported periannular extension, including abscess, pseudoaneurysm and fistula accounted for 37% of left-sided endocarditis. [9–11] In our study, 15.6% (80/512) of patients with aortic valve endocarditis suffered from periannular abscess and destruction of aortic annulus.

Complex infection of the aortic valve with aortic root involvement and destruction of aortic annulus remains a grave challenge to the cardiac surgeon. Patients frequently present profoundly unwell and extensive surgery may be required to correct the underlying anatomical deficits and provide source control in septic patients. Complex aortic valve endocarditis with aortic root involvement and destruction of aortic annulus continues to have a high in-hospital mortality rate of 10 to 40%. [12–15] Our study showed that destruction of the aortic annulus increases mortality and health care costs, and destruction of the aortic annulus is associated with in-hospital mortality and 1-year mortality following cardiac surgery. Improvement of surgical and perfusion techniques can decrease aortic cross-clamp time, cardiopulmonary bypass time, intubation time, ICU retention time, hospitalized time postoperative, paravalvular leak, chest drainage, fresh-frozen plasma, and packed red cells, therefor, contribute to better clinical outcomes.

### Prevention of destruction of the aortic annulus

Preoperative LVEDD, vegetation length, preoperative aortic insufficiency, and serum creatinine before surgery are all parameter of severity of aortic valve endocarditis. Our present study showed that vegetation length and preoperative mitral insufficiency are associated with destruction of the aortic annulus in patients with aortic valve endocarditis. (Table 5) Therefore, early diagnosis and treatment of IE are important. Early surgical intervention is advocated, as aortic valve endocarditis is a progressive and life-threatening disease, and patients with a poor preoperative functional class are at the highest risk for in-hospital death. Early diagnosis and treatment maybe prevent the development of periannular abscess and destruction of aortic annulus.

### Management of destruction of the aortic annulus

Surgical treatment of aortic valve endocarditis with aortic periannular abscess and destruction of the aortic annulus is still a challenging issue with high mortality and morbidity rate in the current era. Surgical method such as aortic valve replacement and aortic root replacement or reconstruction depend on the integrity of aortic annulus. According to the guideline of the Society of Thoracic Surgeons Clinical Practice, if the aortic annulus is healthy and strong enough for suture, it is reasonable to use a mechanical or stented tissue valve following radical debridement completed. If the aortic annulus is disrupted, aortic root replacement or reconstruction is usually the preferred choice of surgery. [16, 17]

There is more and more evidence to indicate that aortic root replacement is associated with advantageous reoperation rates and rate of early paravalvular leak, a hypothesis that is also intuitive and backed by well-established surgical principles. Patients treated with an aortic root replacement were at a lower risk of reoperation and rate of early paravalvular leak at 1-year follow-up. Aortic root replacement accomplishes more comprehensive removal of infected tissue and reconstruction of cardiac morphology compared with aortic valve replacement and therefor can attain lower rates of reinfection and graft deterioration. The aortic root replacement technique is related to an advantageous postoperative profile compared with aortic valve replacement. Aortic root replacement could be recommended as the best practice choice for aortic valve endocarditis with periannular abscess and destruction of the aortic annulus. [18–20]

### Perspectives

In our study, only 54.2% (512/944) patients with aortic valve endocarditis underwent surgery, and 28.8% (272/944) were excluded from surgery because of multiorgan failure at admission. Time between symptoms and admission is associated with multiorgan failure at admission. Vegetation length, aortic insufficiency, and symptomatic neurological complications are parameter of severity of disease, and time between symptoms and admission, vegetation length, aortic insufficiency, and symptomatic neurological complications in group of multiorgan failure at admission were significantly higher than those in group of non-multiorgan failure. Therefore, early diagnosis and treatment of aortic valve endocarditis are important. Early surgical intervention is advocated, as aortic valve endocarditis is a progressive and life-threatening disease. Reaching a rapid and accurate diagnosis in cases of aortic valve endocarditis is still a central challenge of the disease. Delayed diagnosis and initiation of therapy lead to complications and worse clinical outcomes. Because of lack of excellent network of primary, secondary, and tertiary preventions, these patients therefore present late in the hospital and are usually diagnosed and transferred to our tertiary hospital late. A conceptual framework comprising of required baseline information and requirements for executing primary, secondary, and tertiary preventions has been advocated as a best model for aortic valve endocarditis control.

### Strengths and limitations of the study

Our study for the first time clarified the risk factors, significance, and management of destruction of the aortic annulus in aortic valve endocarditis, adding value to the existing literature. Limitations of the present study include its retrospective design. There may be a selection bias because of the retrospective nature of the study and our hospital as a referred center. Well-designed research such as prospective cohort studies are needed and programs aiming at the reduction of in-hospital morbidity and mortality caused by aortic valve endocarditis are encouraged.

Conclusions: In our study, destruction of the aortic annulus increases mortality and health care costs. Optimization of pre-, peri-, and postoperative factors can reduce mortality and morbidity in aortic valve endocarditis. Aortic root replacement could be recommended as the best practice choice for aortic valve endocarditis with periannular abscess and destruction of the aortic annulus.

## Data Availability

yes

## Declarations Ethical Approval

The experiment protocol for involving humans was in accordance to Helsinki Statement and national guidelines and was approved by the Medical Ethics Committee of The People’s Hospital of Guangxi Zhuang Autonomous Region, and They gave the authors approval to waive the need for patient consent for publishing data in the study about the patients.

## Acknowledgements

We thank doctor Li and Long for their help.

